# The Impact of Craniotomy and Surgical Fixation Devices on the Efficacy of Tumor Treating Fields in Glioblastoma Treatment

**DOI:** 10.64898/2025.12.10.25342010

**Authors:** Fang Cao, Nikola Mikic, Konstantin Weise, Axel Thielscher, Anders R Korshøj

## Abstract

Glioblastoma is increasingly treated with Tumor Treating Fields (TTFields), but how post-craniotomy anatomy and fixation hardware alter delivered fields is unclear. We used finite-element modeling in a realistic head model to simulate TTFields after a standard bone flap with either a non-penetrating fixation plate or a penetrating skull clamp, and compared results to an intact-skull baseline across a range of clinically used array layouts. Bone gaps increased mean brain electric-field magnitude by ∼10–20%. Non-penetrating plates caused only minimal, localized changes relative to the bone-gap condition. In contrast, penetrating clamps produced strong but spatially confined increases: local mean fields were ∼6–8x higher within 5–10 mm of the device, with ≥ 50% enhancement extending ∼50–60 mm depending on whether the gap was modeled as healed scalp (soft-tissue-like) or healed bone; this enhancement decayed with distance. These simulations, performed in a single head model with literature-based tissue conductivities, suggest that penetrating hardware can substantially modulate local TTFields delivery, whereas non-penetrating plates have minimal impact. Accounting for post-surgical anatomy and hardware in TTFields planning may improve dose targeting.

## Introduction

Glioblastoma (GBM) is an aggressive primary brain tumor with limited effective therapies and poor prognosis [1]. Tumor Treating Fields (TTFields) deliver low-intensity, intermediate-frequency alternating electric fields (100–500 kHz) via scalp arrays and disrupt mitosis, thereby inhibiting tumor-cell proliferation [2–4]. In the EF-14 randomized trial (NCT00916409), adding TTFields to maintenance temozolomide improved progression-free and overall survival compared with temozolomide alone [5, 6]. Consistent benefits and patterns of use have been described in real-world cohorts [7], and these data underpinned FDA approval of TTFields with temozolomide for newly diagnosed GBM in 2015 [8, 9]. Recent reviews and post-marketing surveillance summarize advances in mechanisms, clinical practice, and the global safety profile in more than 25,000 patients with CNS malignancies [10, 11]. In parallel, TTFields dosimetry and treatment-planning studies highlight opportunities to optimize delivery and refine dose–response relationships [12].

TTFields is typically initiated after craniotomy, where a bone flap is replaced and secured with fixation hardware. The residual bone gap and the electrical properties and geometry of the hardware may alter current paths and local field intensities in the brain. Consistent with this, skull defects (craniectomy) have been shown to markedly modulate TTFields distributions [13, 14]. In particular, devices that penetrate the skull can plausibly influence dose near target tissue [15]. Beyond cranial fixation hardware, a recent case report describes safe TTFields use in a patient with an implantable cardiac pacemaker, suggesting that interactions with certain cardiac implantable electronic devices (CIEDs) may be clinically manageable with appropriate precautions [16].

To examine these effects, we consider two classes of cranial fixation hardware that differ in whether metal penetrates the skull: a non-penetrating plate (e.g., CranioPlate) and a penetrating double-sided clamp (e.g., Craniofix). The analysis aims to identify mechanistic principles that extend beyond the individual devices to the broader classes they represent.

Our study asks how the craniotomy gap and fixation hardware modify the spatial distribution and magnitude of TTFields within gray matter, white matter, peri-tumor, and residual tumor. Using finite-element simulations in a realistic head model, we compare an intact skull, craniotomy alone, and craniotomy with each device under clinically used array layouts. We report mean and peak field intensities and localized differences relative to the intact-skull baseline.

## Methods

### Ethical Considerations and Consent

This study involves the use of human subject data from the OptimalTTF-2 trial (NCT04223999), see also Mikic et al., 2024 for final clinical results [17], which complies with the international guidelines for Good Clinical Practice (ICH-GCP) and DS/EN ISO 14155:2012. The trial received approval from the Central Denmark Region Committee of Health Research Ethics, The Danish Data Protection Agency, and the Danish Health Authorities. In accordance with The Danish Act on Processing of Personal Data, we ensured rigorous data protection and privacy measures. Written informed consent was obtained from all participants or their legal guardians, authorizing their participation in the trial and the use of their non-identifiable imaging data for future in silico studies. This consent process was designed to protect participants’ privacy and to comply with ethical standards governing human research.

### Head Model Generation

To investigate the impact of cranial fixation devices on the electric fields generated by TTFields, a detailed head model was constructed using structural MR images and the Finite-Element Method (FEM) [18]. The “Ernie” dataset provided by the SimNIBS software package was utilized for the proof-of-concept demonstration performed in this study [19].

#### Dataset and Initial Segmentation

The head model was based on the “Ernie” dataset from SimNIBS. Tissue segmentation was automated using the CHARM pipeline [20] with T1 and T2 weighted images from this dataset, identifying eight tissue compartments. The model also included other tissue compartments such as white matter, gray matter, CSF, scalp, skull, muscle, blood, and eyeballs.

#### Creation of Virtual Tumor Resection Cavity

Visual inspection allowed manual configuration of a virtual tumor resection cavity based on postoperative MRI and CT scans [21]. A sphere-shaped tumor resection cavity and a cylindrical funnel structure were introduced, along with a sphere-shaped tumor remnant beneath the cavity (Fig 1).

**Fig 1.**
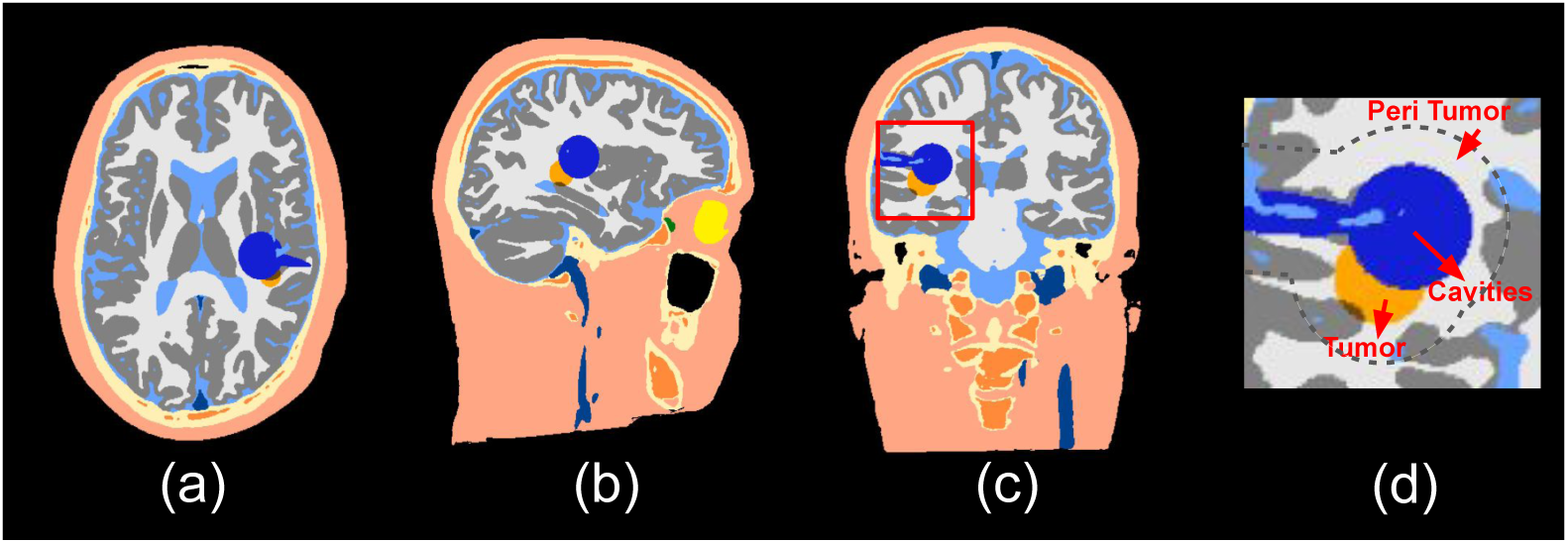
The modified head model derived from the MRI of a healthy individual (A axial, B sagittal, C coronal orientations and D zoomed-in view of C highlighting the tumor/cavity/peri-tumor regions). To resemble a postoperative MRI of an actual patient, specific pathology features were incorporated: a sphere-shaped tumor resection cavity of 2.5 cm diameter was introduced, accompanied by a cylindrical funnel structure of 0.8 cm diameter on top, simulating the surgical entry to the tumor. Beneath this cavity, a sphere-shaped tumor remnant of 2.5 cm diameter was added.

#### Bone Gap and Fixation Devices

To mimic brain surgery, a bone flap was added to the skull model. The bone flap had a diameter of 75.4 mm, and the width of the gap between the bone flap and the skull was 2 mm, reflecting a real craniotomy scenario. We incorporated two types of cranial fixation devices: the SterileTrac long dog bone set (referred to as ‘non-penetrating plate’) and Craniofix (referred to as “penetrating clamp”) (Fig 2).

**Fig 2.**
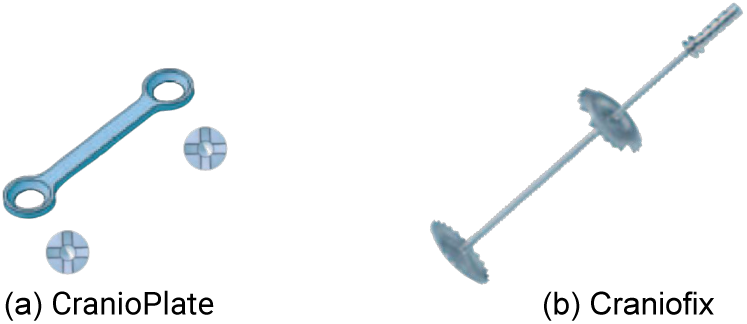
Representative cranial fixation hardware examined in this study: (a) non-penetrating plate (CranioPlate; Biomet Microfixation) and (b) penetrating clamp (Craniofix; Aesculap). Devices are shown as examples only [22].

A. The non-penetrating plate (SterileTrac long dog bone set by Biomet Microfixation): A straight 2-hole fixation device, model FM960T, with a length of 11 mm. Paired with self-drilling screws, model FM921T, each 4 mm in length.
B. The penetrating clamp (Aesculap): A screwless, double-sided fixation system. Each side of this clamp boasts a round cap with a diameter of 11 mm, utilizing a pin-grasping mechanism for secure attachment.

#### Head Model Generation and Conductivity Assignment

After incorporating the fixation plates and the fixation clamps into the three-dimensional voxel segmentation of the head (image resolution 0.5 mm³ isotropic), the updated segmentations were transformed into tetrahedral meshes as needed for the FEM calculations using tools provided by SimNIBS [23–25].

Conductivity values were assigned to various compartments of the head model. These included skin (0.25 S/m), sponge bone (0.025 S/m), compact bone (0.008 S/m), CSF (1.654 S/m), gray matter (GM) (0.276 S/m), white matter (WM) (0.126 S/m), tumor (0.24 S/m), and necrotic tissue (1.0 S/m) [26, 27].

We explored various scenarios involving conductivity within the bone gap, set at 0.25 S/m or 0.008 S/m. For the bone gap, our high-end conductivity value was chosen to mimic the conductivity of the skin, while the low-end value represented that of bone tissue. We made this choice anticipating the formation of scar tissue inside the gap.

Regarding the titanium fixation device, we examined a broad conductivity spectrum, including a high-end value of 1e6 S/m and a low-end value of 1e-6 S/m [28]. This decision was informed by Mercadal et al., who highlighted that metal implants, including titanium, can exhibit insulator-like behavior when subjected to moderate electric fields. This insulator-like behavior is especially pronounced in scenarios characterized by weak surrounding electric fields and a lack of charges around the fixation device, and for direct currents (as opposed to the currents induced by TTFields at ¿ 100 kHz). However, by examining both conductivity extremes, we aimed to fully assess the range of potential impacts of the titanium fixation device on electric field simulations.

### Transducer Array Placement

To analyze the field distribution for a single opposing array pair, we positioned the transducer arrays on the skin surface of the head model. The array pair was rotated in 15° intervals around the central craniocaudal z-axis, a technique frequently used in TTFields dosimetry studies [29–31]. This configuration is depicted in Fig 3. We utilized two 3 × 3 transducer arrays with electrodes measuring 1 mm in height and 2 cm in diameter. The electrodes were spaced at center-to-center distances of 45 × 22 mm.

### TTFields Dosimetry

Based on the established placements of the transducer arrays, we conducted a dosimetry analysis of TTFields, using a baseline-to-peak current strength of 0.9 A as used by the Optune device. While factors such as treatment duration, frequency, and field orientation also play roles in the efficacy of TTFields [32–34], our focus was on the field intensity as the primary metric. This approach aligns with conclusions drawn in previous studies [29, 35–40]. We used the Finite-Element Method (FEM) implemented in SimNIBS v4.0 [41] to determine the electric field distribution [18, 42].

The primary objective of this study was to discern the influences of the bone gap and fixation devices on the electric fields within an artificial setup using the Ernie head model. To achieve this, we analyzed four distinct scenarios, as depicted in Fig 4.

**Fig 3.**
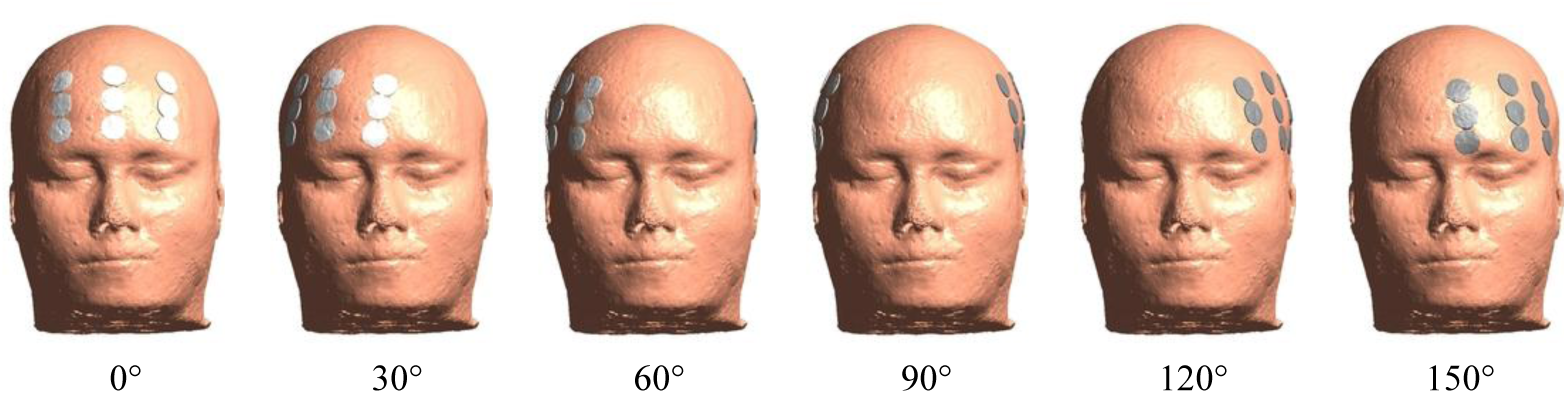
Placement variations of the transducer arrays. The electrode arrays, depicted in white and gray, were positioned in opposing locations on the head model. The figure shows the rotation of the array pair around the central craniocaudal z-axis in 15° intervals. The rotation occurred 5 cm above the central horizontal plane. The subfigure highlights six representative positions at angles of 0°, 30°, 60°, 90°, 120°, and 150°.

**Fig 4.**
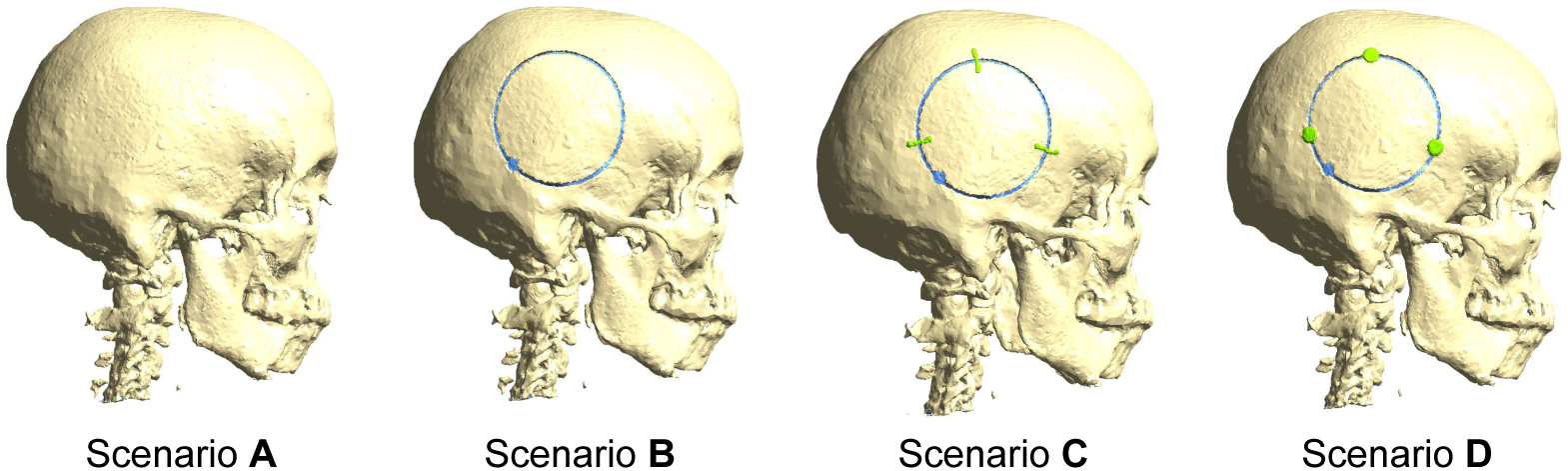
Varied cranial configurations for the TTFields analysis: A. Baseline scenario - Skull model without bone flap or fixation device. B. Bone flap scenario - Skull model with bone flap, without fixation device. C. Plates scenario - Skull model with bone flap and fixation plates. D. Clamps scenario - Skull model with bone flap and fixation clamps.

**A. The Ernie model with untouched skull (Baseline):** Representing the absence of surgical alterations, this configuration provided a control model.
**B. The Ernie model with bone flap and high conductivity gap:** By introducing a bone flap along with a gap of high conductivity between the flap and the skull, this scenario enabled an exploration of how these elements might affect the electric field distribution.
**C. The Ernie model with bone flap and three non-penetrating plates:** Integrating non-penetrating devices, characterized by high conductivity, this model facilitated an investigation into the potential modifications to the electric fields arising from these specific fixation devices.
**D. The Ernie model with bone flap and three penetrating clamps:** Incorporating penetrating devices, also modeled as conductors, this scenario sought to assess their distinctive impacts, allowing for a direct comparison with the non-penetrating plates.

It should be noted that scenarios A and B integrated the cases of bone flaps and fixation devices with low conductivities. We will provide further clarification on this in Section “Overview of Case Studies” of the Results.

By analyzing these diverse configurations, our dosimetry study provided a detailed understanding of how different fixation devices and bone structure alterations might interact with TTFields.

### Analysis of Electric Field Variations in Brain Regions

We evaluated the electric fields in the GM, WM, peri-tumor, and residual tumor regions using the Ernie head model, which were constructed as tetrahedral meshes. Let *E*_base_ denote the electric fields in the baseline scenario and *E*_variant_ represent the electric fields in the comparative scenario. Additionally, let *V* be the volume of a single tetrahedron within the mesh.

To measure the deviation of the comparative scenario from the baseline, we computed the percentage difference using the formula:

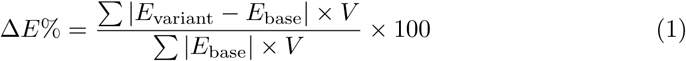

In this formula, the numerator represents the total weighted differences across all tetrahedra, and the denominator serves to normalize these differences based on the baseline electric field.

To highlight the most pronounced electric field in the comparative dataset, we also identified its peak value, which was defined as the 99th percentile of the field intensities.

## Results

### Overview of Case Studies

In our investigation, we analyzed the electric field distribution within the context of varying conductivities in bone gaps and fixation devices. To facilitate our study, we categorized the cases involving bone gaps and fixation devices with low conductivities into scenarios A and B. These categories allowed us to better understand the similarities and differences in electric field behavior.

**Fig 5.**
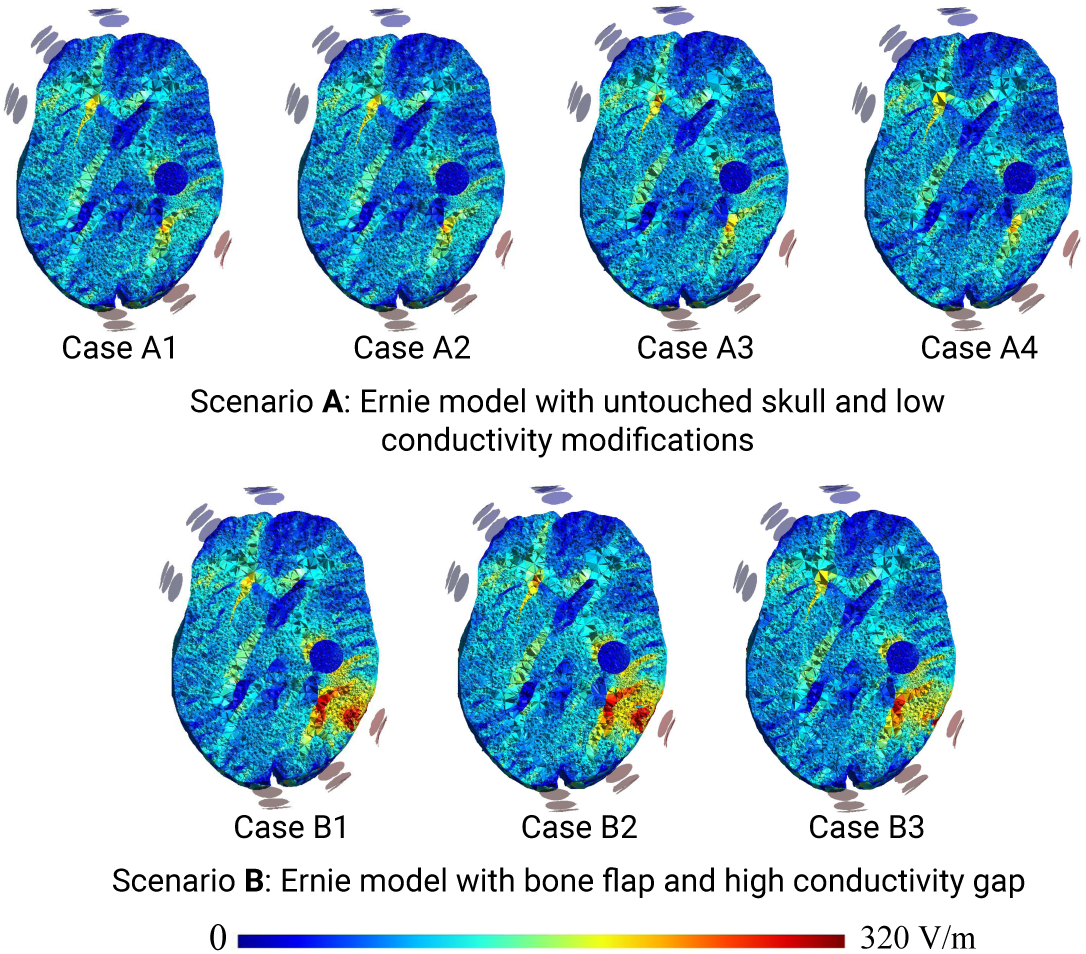
Electric field distribution comparison in scenarios A and B: The first row presents electric field distribution cases within Scenario A, encompassing the baseline case A1 and cases A2-4 with low conductivity bone gap and fixation device modifications. The second row displays electric field distribution cases within Scenario B, featuring a bone flap and a high conductivity gap (B1). Additionally, on top of B1, low conductivity fixation devices are introduced in cases B2-3.

#### Scenario A: Ernie Model With Untouched Skull (Baseline)

In Scenario A, we focused on the Ernie model with an untouched skull, which served as the baseline for our investigation. Within this scenario, we explored various cases, each representing different configurations and conductivity attributes. Fig 5A illustrates the baseline case (A1) and the associated cases A2-4. These cases consistently displayed nearly identical electric field outcomes, highlighting their fundamental similarities.

**Case A1:** Case A1 served as a direct reflection of Scenario A, featuring the Ernie model with an untouched skull. This case established the baseline for comparison.

**Case A2:** Similar to Case A1 but included a bone gap with low conductivity.

**Case A3:** Introduced a bone-shaped fixation plate with low conductivity.

**Case A4:** Mirrored Case A3 but featured a fixation clamp with low conductivity.

#### Scenario B: Ernie Model With Bone Flap and High Conductivity Gap

Scenario B investigated the electric field distribution within the Ernie model, now with the inclusion of a bone flap and a high conductivity gap, representing a surgical alteration. Fig 5B visually represents this bone gap scenario, including cases B1-3, which share similar electric field behavior.

**Case B1:** Case B1 directly represented Scenario B, introducing a bone flap and a high conductivity gap, thus altering the electric field distribution.

**Case B2:** Similar to Case B1, we maintained the high conductivity gap but integrated the non-penetrating plate with low conductivity.

**Case B3:** Incorporated the penetrating clamp with low conductivity into the bone gap scenario.

This analysis enabled us to comprehensively examine electric field behavior in Scenario A and B under various conditions, providing valuable insights into the impact of different configurations and conductivities on the electric field distribution.

In the forthcoming sections, our analysis will specifically focus on examining electric field interactions in the presence of bone gaps and fixation devices. To facilitate this analysis, we will designate Case A1 as the reference point for baseline Scenario A and Case B1 as the reference point for bone gap Scenario B. Furthermore, to explore the impact of high conductivity attributes, we will investigate two additional scenarios:

**Scenario C:** In this scenario, we maintain the Ernie model with a bone gap and set both the bone gap and non-penetrating plate conductivities to high.

**Scenario D:** Similarly, in this scenario, the Ernie model includes a bone gap, and we set both the bone gap and penetrating clamp conductivities to high.

### Comparison of Electric Field Distributions: Assessing the Impact of Bone Gap and Fixation Devices

In this part of our investigation, we utilized the Ernie head model to investigate the electric field distributions under various conditions, including the presence of bone gaps and different fixation devices. Fig 6 offers a detailed visualization of these scenarios, showing the electric field patterns across different electrode patch layouts.

The bone gap, having a width of 2 mm, had a certain influence on the electric fields. When electrodes were placed over this gap, there was a potential for an elevation in the electric field beneath it. This elevation, however, was observed to be limited and did not lead to a substantial shift in the overall electric field distribution within the Ernie head model.

The non-penetrating plate, shaped like a bone, resulted in a localized increase in the electric field compared to the bone gap scenario. However, when considering its overall impact on the electric field distribution, it was comparable to the scenario with a bone gap alone. Consequently, we can conclude that the influence of the non-penetrating plates on the electric field was minimal.

The results generally showed no significant variation in most parts of the brain. However, a notable exception was observed when electrodes were placed near the fixation clamp. This device could significantly alter the electric field when an electrode patch was positioned close to it. This effect was likely due to its double-sided shape, which allows the metal to penetrate deeper beneath the skull. The most intense electric fields were often produced when the edge of an electrode was directly above the fixation clamp. Even when electrodes were placed at a greater distance from the clamp, such as in the 0° scenario, these devices continued to influence the electric fields.

To better understand the intricate link between cranial alterations and TTFields, we used three metrics to measure electric field variations: the weighted mean, peak values, and percentage differences compared to the baseline case. This analysis aimed to pinpoint the effects and contributing factors of the bone gap and the two fixation devices, the non-penetrating plate and the penetrating clamp, on the electric fields. The findings provide insights into how these surgical procedures could influence electric fields, particularly in critical areas such as the tumor and resection cavity.

Fig 7 shows the electric field intensities across various brain regions. We observe that the electric fields experience a roughly 10-20% increase due to the creation of the bone gap during craniotomy. This increase is typically less than 20% in areas like the white matter, gray matter, peri-tumor, and residual tumor. While our preliminary results indicate that the impact of the bone gap is relatively limited, it is important to keep these variations in mind for future clinical settings.

**Fig 6.**
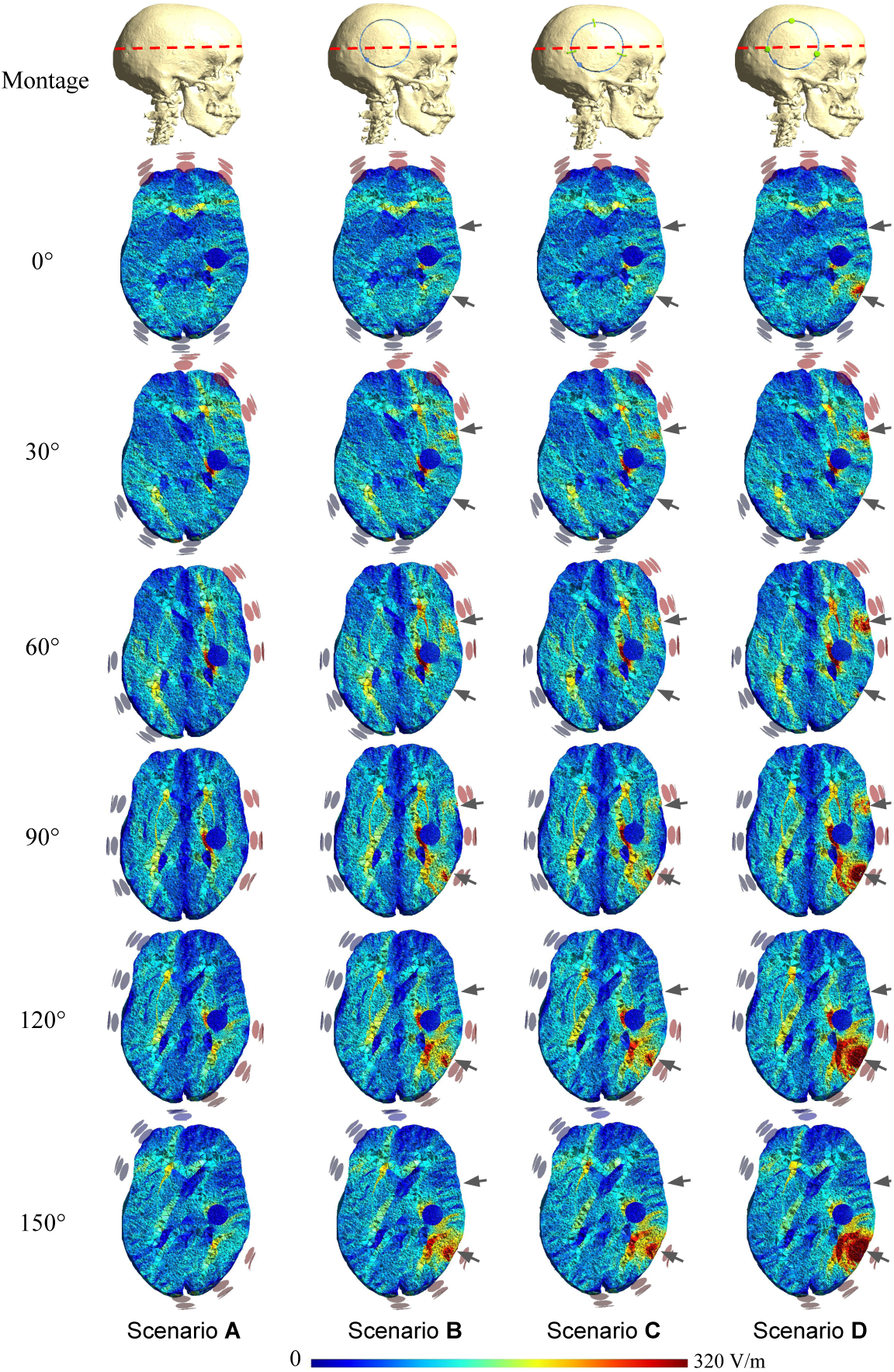
Electric field distributions across different scenarios and electrode patch layouts in the Ernie head model. The four columns correspond to the four distinct scenarios: (1) Scenario A, the head model with an untouched skull (baseline); (2) Scenario B, the head model with a bone flap; (3) Scenario C, the head model with a bone flap and three non-penetrating plates; and (4) Scenario D, the head model with a bone flap and three penetrating clamps. The first row indicates the axial view position in the head model. The subsequent six rows represent electrode patch rotations at 0°, 30°, 60°, 90°, 120°, and 150°. Arrows within the image indicate the locations of the bone gap and the fixation devices in the axial view. All colors have been standardized for consistency.

**Fig 7.**
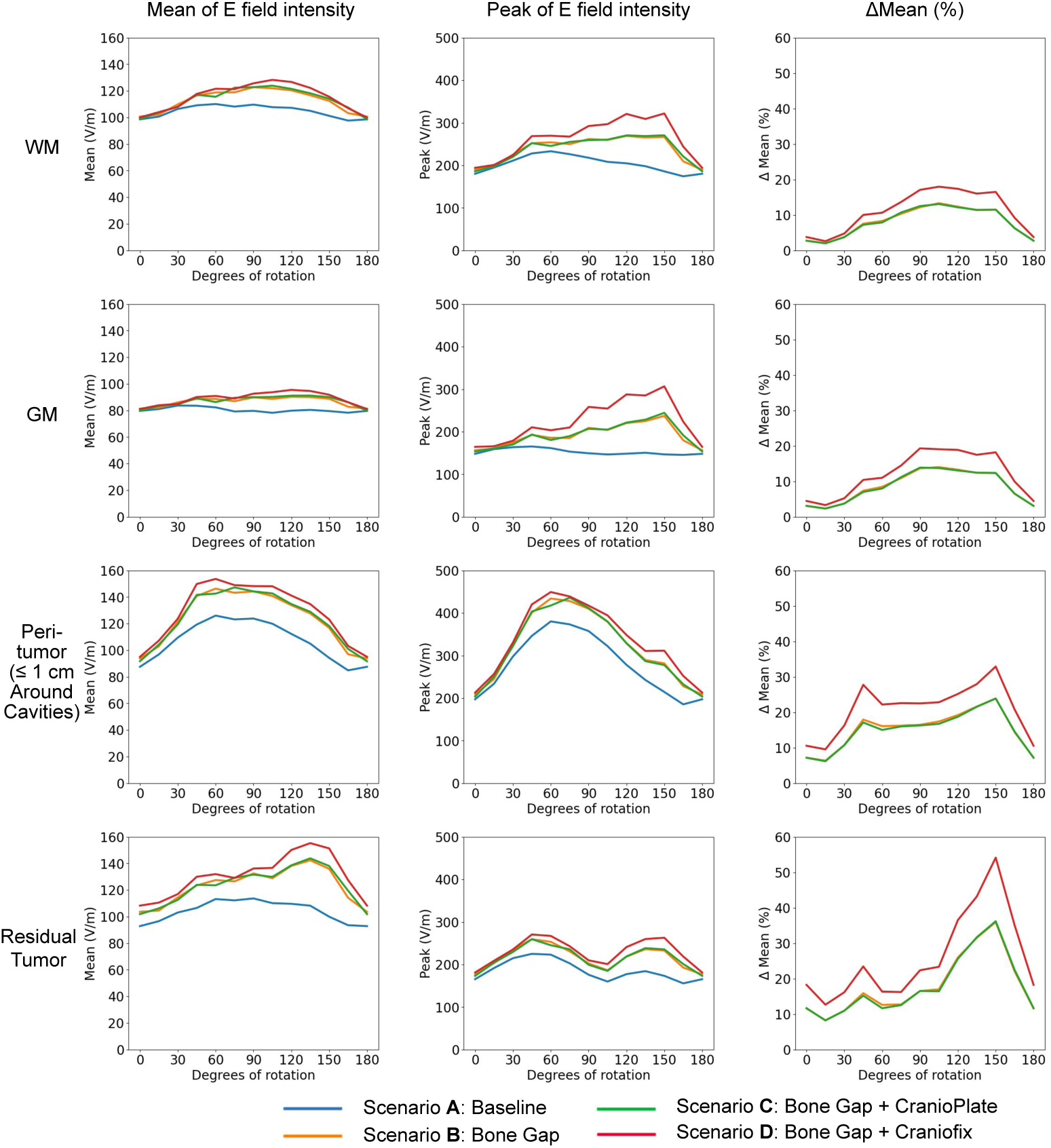
Comparative analysis of TTFields intensities across brain regions under scenarios A-D. This illustration depicts TTFields intensities in white matter, gray matter, the vicinity of the resection cavity, and the residual tumor across four scenarios: baseline (blue), bone gap (yellow), bone gap with the non-penetrating plate (green), and bone gap with the penetrating clamp (red). The x-axis represents rotation degrees around the central craniocaudal z-axis, and the y-axis denotes field strength. The first two columns display mean and peak field intensities, while the last column shows relative enhancements (percentage difference) in field intensities when introducing the bone gap and fixation devices compared to the baseline.

Regarding the non-penetrating plate, the findings confirm our previous analysis. The device introduces only a minimal and localized change in the electric field when compared to the bone gap scenario. In the Ernie head model, this change is almost undetectable, suggesting that its therapeutic influence is likely negligible.

In the presence of the penetrating clamp, we generally observe a difference of 5-10% in electric field intensities relative to the baseline scenario. However, under certain transducer orientations, regions like the peri-tumor and the residual tumor might see electric field increases of up to 40% and occasionally surpassing 50% compared to the baseline. A detailed analysis of the penetrating clamp’s impact will be covered in the following subsection.

### Effects of the Increasing Electric Fields Underneath the Penetrating Fixation Clamp

In this section, we investigate the influence of the penetrating clamp on electric field intensities. We refer to our previously established scenarios A to D and introduce a new case involving a single fixation clamp without a bone gap. In this setup, electrodes are positioned directly above the clamp, simulating a post-craniotomy situation where the bone gap has completely healed. This approach ensures that any observed changes are solely attributable to the penetrating clamp, eliminating potential electric field increases resulting from the bone gap. Consequently, this comparison allows us to assess the minimal electric field change that the penetrating clamp may introduce.

Fig 8 provides a visual representation of electric field variations influenced by the penetrating clamp. We observe a noticeable increase in electric field intensity near the clamp. Specifically, we identify an amplification of over 150% in the electric field around this device. The enhanced electric field region is approximately sphere-shaped, centered around the penetrating clamp, and its dimension is larger than a single TTFields treatment electrode (here 20 mm in diameter). The significant electric field increase within this region highlights the substantial impact of the penetrating clamp on the electric field distribution.

Our analysis, further illustrated in Fig 9, examines the increase in electric field intensity due to the penetrating clamp. The data show that, compared to the baseline scenario, the mean and peak values, as well as the percentage enhancement of the electric field, generally rise within spherical regions centered on the device. These regions range from a minimal 5 mm radius to a maximum of 170 mm, covering the entire brain. The most substantial increase in intensity occurs within a 50 mm radius of the penetrating clamp, which is particularly relevant for treatment efficacy near pathological areas. Within this key radius, the intensity of the electric fields surges from 780% at the smallest radius to 56% at 50 mm. Beyond this 50 mm mark, the increase in electric field intensity drops below 50%, and the increase stabilizes at 12% beyond a 130 mm radius, a trend that continues up to the full extent of the brain.

The analysis also includes Scenario D, which assesses the effects of a bone gap together with the penetrating clamp. This scenario shows a similar pattern of intensity enhancement but with slight variations in magnitude. In Scenario D, the most significant enhancement occurs within a 60 mm radius around the device, ranging from 625% at a 5 mm radius to 53% at a 60 mm radius. Beyond the 60 mm mark, the increase in intensity falls below 50%, stabilizing at 18% beyond a 130 mm radius. The data indicate that incorporating the bone gap slightly lowers the maximum electric field values, especially within the 5-10 mm radius, resulting in a more diffuse electric field across the brain rather than one concentrated around the penetrating clamp.

**Fig 8.**
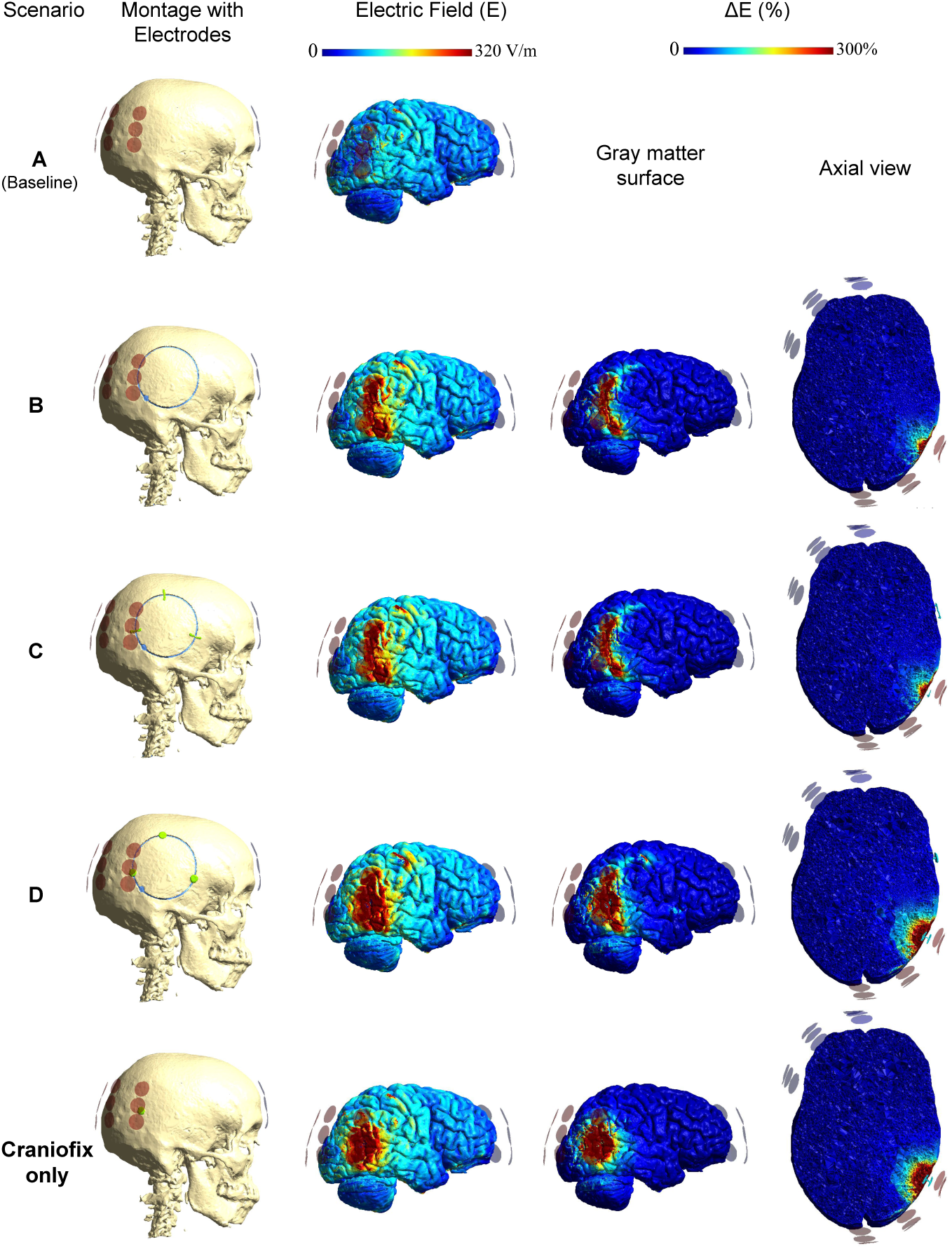
Visualizing the impact of the penetrating clamp on electric fields. Each row corresponds to a unique scenario, and specific columns focus on various aspects of the analysis. Rows 1 to 4 align with scenarios A to D, with Scenario A serving as the baseline reference. Row 5 illustrates a scenario featuring a single fixation clamp, excluding any bone gap. The columns present the montage with electrodes, electric fields on the gray matter surface, percentage differences in the electric field on the gray matter surface, and percentage differences in the electric field in the axial view. All colors have been standardized for consistency.

**Fig 9.**
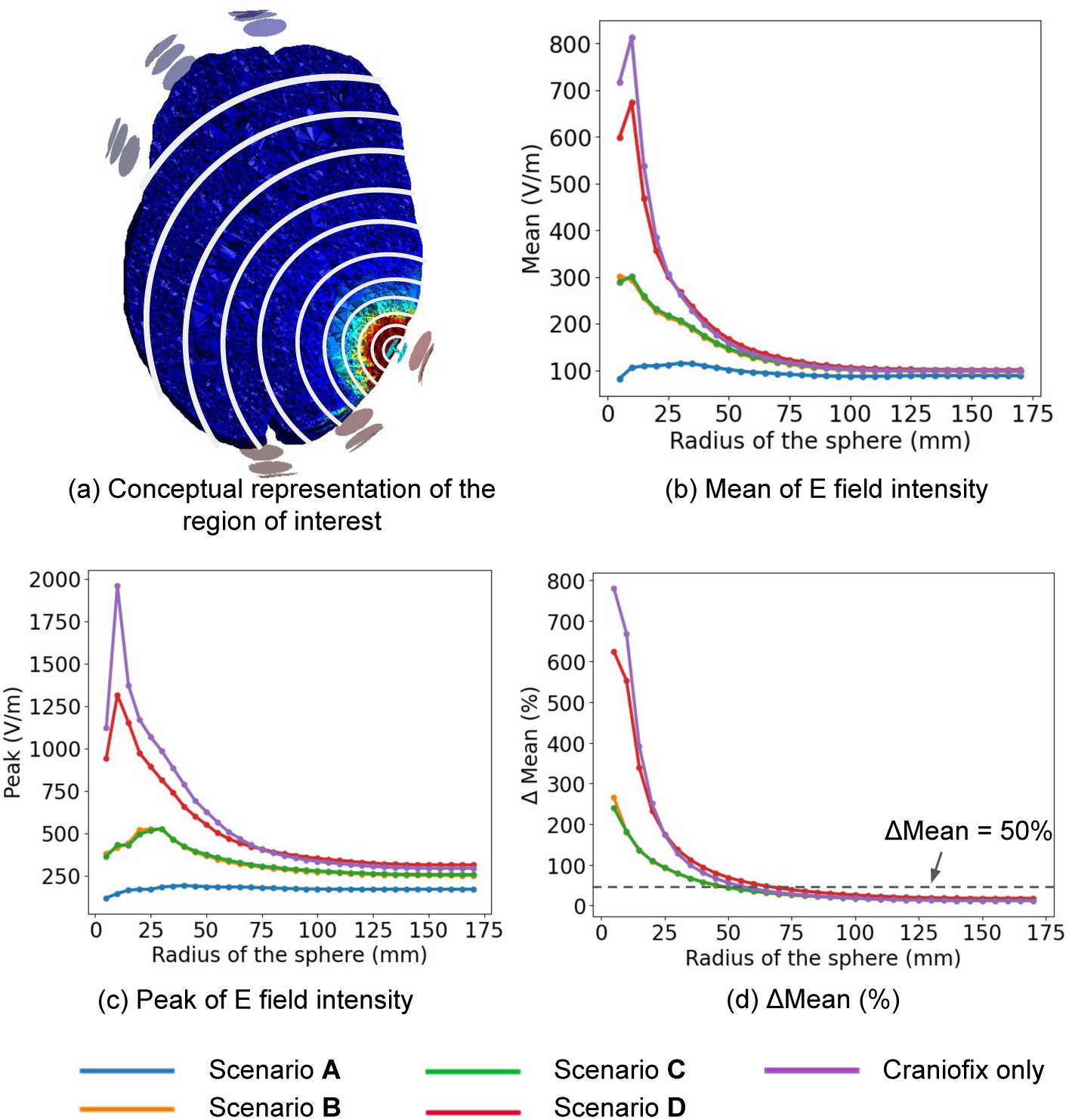
Comparative analysis of TTFields intensities with penetrating clamp. Comparison of the electric field intensities across scenarios A-D and an additional scenario that incorporates a penetrating clamp without a bone gap. (a) defined regions of interest in the brain, employing spheres with radii that increase in 5 mm increments. The spheres range from a minimal radius of 5 mm to a size of 170 mm that encompasses the entire brain, including white matter, gray matter, and tumor regions. (b) and (c) show the weighted mean and peak field intensities for these scenarios. (d) plots the percentage difference, quantifying the change in the electric field intensity due to the various surgical interventions and fixation devices, comparing each scenario to the baseline. All graphs are plotted with the sphere’s radius measured in millimeters, on the x-axis.

The findings from scenario B, which features only a bone gap, suggest that the gap alone does not provide as significant an enhancement in electric field intensity as the scenarios that include the highly conductive penetrating clamp. In this scenario, the electric field enhancement ranges from 266% at a 5 mm radius to 57% at a 40 mm radius. After the 40 mm radius, the enhancement continues to decrease, reaching 12% at a 130 mm radius and then stabilizing at this level across the entire brain. Additionally, the results from scenario C, which has both a bone gap and non-penetrating plates, confirm that plates do not significantly influence the electric field intensities within the brain.

Our research clearly demonstrates that the penetrating clamp significantly impacts the electric field intensity in the brain in a localized manner. The data suggest that the strategic placement of electrodes near the device markedly affects the electric field in the region close to the device, potentially enhancing treatment efficacy. Furthermore, the interplay between the bone gap and the device’s presence illustrates the complex dynamics of surgical alterations and their impact on electric field modulation, essential considerations for therapeutic planning and outcomes.

## Discussion

This study investigated the influence of cranial structural modifications, specifically bone gaps and fixation devices, on the efficacy of Tumor Treating Fields (TTFields) therapy. The insights garnered from our research contribute to a deeper understanding of how these variables can be manipulated to improve clinical outcomes [7].

1. **Bone gap impact:** Our simulations indicate that bone gaps resulting from craniotomy procedures can lead to a 10-20% increase in electric fields, especially evident in regions such as white matter, gray matter, peri-tumor, and residual tumor. This increase, though modest, suggests potential clinical implications that should be considered when devising therapeutic strategies. Future studies on TTFields dosimetry should incorporate accurate representations of surgical interventions to enhance treatment planning and efficacy.
2. **Non-penetrating fixation device impact:** The non-penetrating plate, designed with bone-like shapes, showed a minimal and localized change in the electric field within the Ernie head model. Given its limited influence, it’s likely that its impact on TTFields treatment efficacy is clinically negligible. Therefore, non-penetrating plate may not be the optimal choice for enhancing TTFields in clinical settings.
3. 3. **Penetrating fixation device impact:** The penetrating clamp significantly influences electric field strengths. Our studies show that when the clamp acts as a perfect metal conductor, it amplifies current penetration. Specifically, in cases where a post-craniotomy bone gap is modeled as healed scalp tissue, we observe an increase of over 50% in the electric field within a 60 mm radius of the device, with intensity spiking by 625% within 5 mm of the device. When the bone gap has fully healed, the intensity boost reaches up to 780% at the 5 mm distance, which is higher than in the healed skin scenario, and remains over 50% within a smaller 50 mm radius. These findings underscore the potential for customizing device interactions to maximize therapeutic benefits and should be incorporated into future TTFields models.

### Comparative Analysis of Cranial Fixation Devices

While this study primarily focused on one penetrating fixation device (Craniofix), it is important to note that other cranial fixation devices, such as those from Stryker, KLS, and Integra, are likely made from similar materials and have comparable shapes. The key difference between these devices and the penetrating clamp is whether the metal passes through the skull (as with the clamps) or not (as with the plates). Although a detailed comparative analysis of these devices is beyond the scope of this study, future research should consider these variations to provide a comprehensive understanding of their impact on TTFields efficacy. Given emerging data that TTFields can impact spinal metastases as well, the influence of spinal rods, screws, and cages on local field delivery merits dedicated modeling and experimental validation [43].

### Optimization Strategies for TTFields Therapy with Penetrating Fixation Devices

Our findings advocate for a strategic application of the penetrating clamps to substantially enhance TTFields therapy. The recommended approaches for maximizing the therapeutic impact include:

1. Prioritizing the use of penetrating device over non-penetrating plate for bone plate fixation in patients undergoing TTFields therapy post-surgery. Placing the penetrating clamps close to the pathological region can significantly boost the therapeutic electric field. Implementing multiple devices to cover a larger area, while keeping them within a 50-60 mm radius of the pathological region, is likely to enhance efficacy.
2. Positioning one TTFields transducer of each pair near the penetrating clamps and close to the pathological region to amplify the electric field, while placing the other transducer in the pair on an unaffected scalp area away from the clamps to prevent shielding effects.

### Consideration of Patients Without a Skull Flap

In addition to the above findings, it is important to consider the effect of TTFields on patients without a skull flap, which can occur due to reasons such as infection. Previous studies have provided valuable insights in this area. Specifically, in our previous work, we explored the effect of skull removal (craniectomy) on enhancing TTFields efficacy through computer modeling [13, 14]. Another relevant study discusses the safety and feasibility of skull remodeling surgery to optimize TTFields [17, 30]. Additionally, ongoing research supports the notion that skull modifications, including the absence of a skull flap, can significantly influence the distribution and efficacy of TTFields [12].

By incorporating these practices, TTFields therapy can be customized for each patient’s needs, using penetrating fixation devices to enhance tumor targeting. As another safety consideration, a recent case report describes uneventful TTFields use in a patient with an implantable cardiac pacemaker, suggesting the feasibility of TTFields in the presence of some CIEDs when appropriate precautions are taken [16]. More broadly, the overall safety of TTFields in routine practice is supported by global post-marketing surveillance covering more than 25,000 patients with CNS malignancies treated between 2011–2022 [11]. Continuous refinement of treatment strategies, careful patient monitoring, and adaptable protocols are essential for the progress of TTFields therapy and for improving patient outcomes while mitigating side effects.

## Limitations

While our study has provided valuable insights, it is important to acknowledge several limitations. Firstly, our research relied on a single head model derived from a healthy individual. Real-world scenarios can introduce variations in skull thickness, tissue conductivity, and other patient-specific factors that may affect electric field distributions differently. Secondly, our primary conclusion was drawn based on idealized conductivity values assumed for the bone gap and metal fixation devices. The true conductivity of these components in clinical settings remains uncertain, introducing the possibility of variations that could influence our results. Although we considered both high-end and low-end conductivity cases in our analysis, conducting a more comprehensive investigation that accommodates these conductivity variations would further enrich our understanding of this subject.

## Conclusion

In finite-element simulations of a post-craniotomy head, skull defects modestly elevated intracranial TTFields, non-penetrating plates had negligible impact, and penetrating clamps concentrated fields over centimeter scales near the device. These effects arise from altered skull impedance and conductive pathways introduced by hardware, and they depend strongly on electrode proximity—particularly to clamp edges (see Fig 7–9). Together, the results indicate that post-surgical anatomy and fixation hardware can materially reshape local dose delivery. Incorporating patient-specific bone gaps and hardware into TTFields planning models may therefore improve targeting and risk assessment. Given the single-subject model and uncertainty in frequency-dependent conductivities, treat these simulations as planning hypotheses until they are validated across anatomies and device geometries, including phantom or in vivo measurements of electric field and temperature.

## Acknowledgments

We extend our deepest gratitude to the Danish Cancer Society, the Independent Research Fund Denmark, and Novocure for their generous support and funding, which were crucial in facilitating this research. The pathology in the head model was meticulously recreated based on a postoperative MRI from a patient participating in the OptimalTTF-2 trial, highlighting the significance of collaborative efforts and the permissions granted by involved participants. Our thanks also go out to the clinical and research staff who played an integral role in the OptimalTTF-2 trial, contributing their expertise and dedication to the advancement of our study.

## Data Availability

The datasets generated and analyzed during the current study are partly derived from publicly available sources and partly collected from participants in the OptimalTTF-2 clinical trial under specific ethical approval. The “Ernie” head model, which is utilized in this research, is publicly accessible at the SimNIBS dataset repository (https://simnibs.github.io/simnibs/build/html/dataset.html). All other data generated or analyzed during this study, which are not publicly available due to privacy or ethical restrictions, are available from the corresponding author upon reasonable request and with permission of the Central Denmark Region Committee of Health Research Ethics.

## Author contributions statement

The contributions of the authors to the manuscript are as follows: The conception and design of the study were collaboratively undertaken by all authors. Data collection and analysis, as well as statistical analysis, were carried out by F.C., A.T., and A.R.K. The first draft of the manuscript was prepared by F.C., with all authors contributing to the editing and critical revision of the manuscript to ensure its accuracy and integrity.

Supervision of the project was provided by A.R.K. and A.T. The creation of figures was a joint effort by F.C., A.T., and A.R.K. Lastly, the final version of the manuscript received approval from all authors, confirming their accountability for all aspects of the work.

